# Gender disparities in international COVID-19 clinical trial leadership

**DOI:** 10.1101/2020.08.02.20166751

**Authors:** Muge Cevik, Syed Arefinul Haque, Jennifer Manne-Goehler, Krutika Kuppalli, Paul E. Sax, Maimuna S. Majumder, Chloe Orkin

## Abstract

The COVID-19 pandemic offers considerable possibilities for research and leadership that might equalize opportunity in a new field; however, our study finds instead that more than two-thirds of investigators leading COVID-19-related clinical studies are predicted to be men. These gender disparities in trial leadership during the pandemic suggest that the structural reproduction of inequalities in research has taken place once again in this new academic field. This indicates that policies are needed to facilitate the identification and implementation of strategies to correct gender bias. The active participation of women, trans and gender-nonconforming individuals are needed in research to drive scientific discovery and innovation as well as to better address health disparities.

## Introduction

In addition to the human and financial loss associated with the novel Coronavirus Disease 2019 (COVID- 19) pandemic, COVID-19 has also had a significant impact on both the personal and professional life of the global workforce, including that of the scientific research community [1-3]. Before COVID-19, women occupied fewer leadership positions, led a fewer funded studies, and applied for and received less grant funding than men when they did apply [4-7]. The employment gap that occurs when women take parental leave impacts the rate of academic advancement and in turn the receipt of institutional support to apply for and secure funding [6, 7]. These imbalances contribute to systemic inequalities that hamper women’s access to and progress in science [2, 7, 8]. A review of the gender distribution of 24 COVID-19 national task forces suggests that many committees are comprised of less than a quarter women, indicating that women’s voices and expertise have been excluded from decision making during this unprecedented public health emergency [9].

For example, emerging data suggest that across all disciplines, despite an increased number of peer-reviewed articles submitted to journals during the pandemic, women have published fewer papers than men thus far this year [10]. This may indicate a similarly reduced involvement of women in research leadership positions and an imbalanced distribution of grants and funding -- important indicators of advancement in a scientist’s academic career [4-7, 10, 11]. Being principal investigator (PI) on a clinical trial is strongly associated with advancement to full professor among women academics in infectious diseases [8].

The COVID-19 pandemic offers numerous opportunities in clinical research. These include trials to assess the safety and efficacy of medical interventions, with protocols in various stages of implementation.

Here, we provide a timely analysis to compare the gender distribution of clinical trial leadership in COVID-19 clinical trials.

## Materials and Methods

We systematically searched https://clinicaltrials.gov/ and retrieved all clinical trials on COVID-19 registered from January 1, 2020 to June 26, 2020 using “COVID” as a keyword. As a comparator group, we have chosen two fields that are not related to emerging infections: breast cancer and type 2 diabetes mellitus (T2DM). We retrieved all clinical trials related to these comparator conditions registered at https://clinicaltrials.gov/ within the aforementioned study period as well as those registered in the preceding year (pre-study period: January 1, 2019 and December 31, 2019). Gender of the investigator was predicted using the genderize.io API (application programming interface). This tool has been used to predict the gender of first names in studies regarding gender bias [12, 13] and achieves a minimum accuracy of 82%, with an F1 score of 90% for women and 86% for men [14]. Clinical trials were excluded if i) investigator information was not provided; ii) the genderize.io API could not predict any of the investigators’ gender from their first name; or iii) organization or company names were provided as the investigator. The number of studies that were excluded for the above reasons are reported in the supplementary flow diagram. An exploratory temporal analysis was conducted with the available data. Categorical variables were summarized by frequencies and percentages. We compared groups using Chi-square testing for equality of proportions with continuity correction.[15] The analysis was performed using R (Version 4.0.2). The repository of the datasets used to collect and analyse the data available at https://osf.io/k2r57/.

## Results

We identified 2 345 COVID-19-related clinical trials. Of those, 1 448 had at least one investigator (i.e., principal investigator, study director, or study chair) identified whose gender could be predicted. In the comparator group, we identified 449 trials on breast cancer and 272 on T2DM that were registered. Of those, 274 breast cancer studies and 139 T2DM studies had at least one investigator whose gender could be predicted.

Overall 27.8% of PIs among COVID-19-related studies were predicted to be women, which is significantly different compared to 54.4% and 42.1% for breast cancer (*p*<0.005) and T2DM (*p*<0.005) trials over the same period, respectively (Table 1). While there has been a small increase in the proportion of PIs who were predicted to be women in May 2020, clinical research leadership for COVID-19 among this group was below 25% for the remainder of the study period (Supplementary Material). While 31.4% of study chairs were predicted to be women in COVID-19-related studies, 32.1% (*p*=0.7) and 63.6% (*p*<0.01) were predicted to be women in breast cancer and T2DM trials, respectively. Proportion of study chairs were not significantly different across the three fields.

**Table 1:** Proportion of women leadership in clinical trials between January 1, 2020 and June 26, 2020 and before January 1, 2020.

|  | Jan 1, 2020 - June 26, 2020 |  |  |  |  | before Jan 1, 2020 |  |  |
| --- | --- | --- | --- | --- | --- | --- | --- | --- |
|  | COVID<br>-19 | Breast<br>Cancer | p value | T2DM | p value | Breast<br>Cancer | T2DM | p value |
| **PI** | 27.8% | 54.4% | <0.001 | 42.1% | < 0.001 | 49.7% | 44.4% | 0.158 |
| **Study**<br>**Director** | 28.7% | 48.9% | 0.0103 | 22.2% | 0.751 | 30.5% | 47.6% | 0.028 |
| **Study**<br>**Chair** | 31.4% | 32.1% | 1 | 36.36% | 0.98 | 33.3% | 40.4% | 0.54 |

We also reviewed comparator group studies registered before January 1, 2020 to determine whether the pandemic might have affected gender distribution of trial leadership. We identified 839 clinical trials related to breast cancer and 533 on T2DM over a 12-month period prior to January 1, 2020. Of those, 573 breast cancer studies and 359 T2DM studies yielded at least one investigator whose gender could be predicted. During this “pre-study” period, the proportion of PIs who were predicted to be women were 49.7% and 44.4% for breast cancer and T2DM trials, respectively and the difference was not statistically significant when compared to results from the study period (*p*>0.05).

## Discussion

In this study, we demonstrate that less than one-third of COVID-19-related clinical trials are led by PIs who were predicted to be women, half the proportion observed in non-COVID-19 (breast cancer and T2DM) trials over the same period. The proportion of PIs in breast cancer and T2DM studies also remained similar to the pre-study period. These gender disparities during the pandemic may indicate not only a lack of women’s leadership in international clinical trials and involvement in new projects, but also may reveal imbalances in women’s access to research activities and funding during health emergencies [2, 16].

The COVID-19 pandemic offers numerous opportunities for research and leadership that could equalize opportunity in a new field, but our results suggest the opposite. The pandemic has reinforced the prevailing gender norms in which men continue to be allocated a disproportionate share of the funding, as well as leadership and authorship roles [9, 10, 16]. One potential contributor for this discrepancy is the speed demanded by the research agenda during the pandemic. The sense of urgency in starting clinical trials may lead to an abandonment of any checks and balances around equality and inclusion that would have otherwise encouraged the involvement of women scientists. Many women scientists have already raised concerns about institutional funding distribution lacking gender balance or being left out of research activities despite their expertise [2, 16]. During COVID-19 pandemic, a UK study showed that women were more than twice as likely to take on childcare and schooling responsibilities of children than men, while male academic counterparts leverage professional relationships and networks more effectively [1, 2, 16].

As a community, we must recognise that there is a tendency to “turn to men” in times of crisis both for leadership and scientific expertise [2, 3, 16, 17], highlighting the need to challenge this culture. Research and academia are already competitive; being in the central decision-making group is often challenging due to gender norms, along with roles and rules on how these groups are established and maintained; during health emergencies, these same authoritative circles become more difficult for women scientists to join [2, 16]. Our findings suggest that there is a need for transparency in opportunities and funding that requires actively identifying and addressing the structurally implicit and unconscious biases that favour men.

Our analysis has some limitations. We could include only ∼50-75% of trials for which an investigator’s gender could be algorithmically predicted. Furthermore, while such algorithms allow for the rapid analysis of gender disparities such as those conducted here, they can also be exclusionary to gender non-conforming, non-binary, and trans individuals. Because of this, we have taken care to use the term “predicted gender” throughout the text whenever discussing algorithm-based gender assignment in acknowledgement of gender non-conforming, non-binary, and trans investigators. Beyond these limitations, we did not consider studies that received private funding, which may not have been registered on clinicaltrials.gov; however, it is worth noting that clinicaltrials.gov is an international database with widespread international representation.

In summary, while the COVID-19 pandemic has thus far provided many new opportunities for research, with numerous clinical trials initiated worldwide, a disproportionate proportion of PIs leading COVID-19 related studies are predicted to be men, despite women accounting for 70% of the global health workforce [16]. Our demonstration of gender differences in trial leadership and grant allocation argue for revised policies and strategies that encourage the participation of women in pandemic research. Not only can these women drive discovery and innovation, but they can act to address health disparities and provide role models for the next generation of women scientists [2, 16, 18, 19]. Ensuring gender representation would also reflect the commitment of the global community to promoting gender equality in academic medicine and research: inclusion, diversity, representation, progression, and success for all. A health emergency is not an excuse for disregarding the importance of equitable representation and given the disproportionate burden of such outbreaks for communities who are marginalized due to their gender, sexuality, class, ethnicity, and ability an even greater reason we must advocate for it [20-22].

## Data Availability

The repository of the datasets used to collect and analyse the data available at https://osf.io/k2r57/.

## Authors contributions

MC: conceptualisation, methodology, investigation, literature review, data curation, writing – original draft. SH and MM: investigation, data curation, formal analysis, writing – review and editing; JS, KK, PS: methodology, writing – review and editing, supervision. CO: conceptualisation, methodology, investigation, literature review, writing – original draft, supervision.

## Supplementary material

Flow diagram of process of selection

Gender distribution over time (months)

